# Impact of Dental Disorders on Self-rated dental Health Status of the Elderly in Selected Rural Communities in Kenya

**DOI:** 10.1101/2024.05.28.24308040

**Authors:** Walter Ogutu Amulla, Fletcher Njororai

## Abstract

Low prioritization of oral care for the elderly and inadequacy of resources results in high prevalence of dental disorders in this population in most African countries. This study aimed at assessing the impact of self-rated dental health among the elderly in Kenya. A cross-sectional quantitative study was conducted among 300 subjects in parts of Karachuonyo sub-county of Homa Bay County, Kenya. The sample size was determined using Yamane’s formula based on a study population of 1159. Data was collected through interviewer-administered questionnaires. Majority (64%) of the respondents were aged 65-74 years. Female respondents were more (55.3%) than males (44.75). Further, 8 in 10 of the study sample never had any formal employment with nearly the same proportion (79.3%) self-rating their economic status as poor. Nearly 7 out of 10 (67%) respondents had lost at least one tooth due to medical extraction whereas half of the respondents (52.3%) were having at least one carious tooth at the time of data collection. Tooth loss had the strongest impact on self-rated dental health (X^2^ =27.818, p<0.00001, φ = 0.305) followed by tooth mobility (X^2^ =27.180, *p*<0.00001, φ = 0.301), gingival bleeding (X^2^ =19.378, p=0.000011, φ = 0.254) and cavities (X^2^ =17.757, p=0.000025, φ = 0.243). The study established that dental disorders significantly but disproportionately impacted self-rated dental health of the elderly, with tooth loss being the leading disorder. Increasing provision of services for the elderly for dental health needs is critical in Kenya as in most African countries.

## Introduction

The proportion of elderly population continues to rise globally with a remarkable shift of the increases towards low- and middle-income countries [1]. This demographic transition necessitates a shift of focus for public health systems to r identify and address the needs of this growing elderly population. Whereas in high income countries (HICs) this is well addressed in many ways, for low- and middle-income countries (LMICs), the ongoing demographic transition has largely led to increased morbidity in this population segment [2]. Unmet health needs affect the quality of life as well as life expectancy for the older adults, many of whom may be suffering multi-morbidities [3, 4]. According to the World Health Organization [5], around 70% of sub-Saharan Africa countries spent less than US$ 1 per person per year on treatment costs for oral health care in 2019, the latest year for which data is available.

In literature, there is no universal cut-off age for the elderly. Although both the United Nations and the WHO recommend 60 years plus, others use lower age since the cut-off depends on life expectancy [4, 6]. The Kenya bureau of statistics (KNBS) characterized those aged 65 plus years as elderly [7]. Therefore this study used 65 years and above as constituting the elderly population.

Among the leading morbidities that the elderly in LMICs have had to contend with are oral and dental disorders. These disorders, largely of teeth and gums, include dental caries, periodontal disease, dental trauma, dental pain, tooth mobility and tooth loss [8]. Previous studies across the globe have reported varying but high prevalence of these dental disorders among the elderly. Agrawal et al. [9] reported a dental caries prevalence of 41.9% among elderly Indians whereas Kohli et al. [10], reported a tooth loss prevalence of 35% among the elderly in Oregon, United States. Additionally, a survey conducted in China reported varying prevalence of periodontal disease with gum bleeding prevalence at 88% and calculus at over 95% among persons aged 65-74 years [11].

The African elderly population is particularly affected by poor dental health for various reasons. Low prioritization of oral care for the elderly and inadequacy of resources has resulted in dental disorders being more prevalent among the elderly in most African countries compared with other age-groups, yet there are very few preventive care programs for them and/or tailored for their age-specific needs [12, 13, 14, 15]. For instance, a study among elderly dental patients in Japan reported tooth mobility prevalence of 17% [16], whereas a related study in Sudan reported tooth mobility prevalence of 91% among cases and 53% among controls [12]. Additionally, while most elderly people live in rural areas as most of the Kenyan population, 85%, live in rural areas, dental facilities are mainly found in urban centers and, as opposed to HICs, few social security and retirement programs exist in Africa for health insurance [14, 17]. Also, most of the elderly people may not have worked in formal employment or even worked at all in paid employment all their lives, and many are living in abject poverty [18]. Many of the elderly will also be living alone, after losing their spouse and the generational differences marginalizes them from the rest of the household which exacerbates their health needs in general [19, 20].

In Kenya, as in other African countries, dental disorders remain a leading public health concern among the elderly particularly in rural areas. A study by Crouch and colleagues [21] reported that all (100%) of the elderly people above 65years had experienced dental pain in the past with 83% of the participants suffering dental pain at the time of data collection. Further, a national survey on oral health in Kenya reported that nearly all (99.9%) the adult participants had experienced one form of dental-related disorder in the previous one year [22]. Another study conducted in parts of western Kenya reported that nearly 9 in 10 (86.3%) elderly persons had lost six lower front teeth due to a cultural custom [23].

Furthermore, dental disorders have been documented to adversely impact the health and wellbeing of the elderly. A study conducted in southern Brazil reported that toothache affected sleeping, talking, cleaning of teeth, eating and emotional state among elderly persons [24]. Additionally, complete tooth loss (edentulism) has been reported to cause cognitive and physical impairment in among the elderly and associated with reduced longevity in several studies [25, 26]. Dental disorders have also been reported to compromise food intake leading to malnutrition with its sequelae of systemic diseases [27]. A meta-analysis by Toniazzo and others [28] in 2018 associated edentulism with poor nutrition among the elderly across several studies.

Moreover, dental diseases share several risk factors with other noncommunicable diseases including excess sugar consumption, tobacco use and harmful use of alcohol hence their prevalence in the population often signify risk of multimorbidity [29]. A global study involving 39 countries reported that the loss teeth predicted cardiovascular outcomes including cardiovascular death and stroke [30]. Low health literacy surrounding oral hygiene, and dental health in particular, also contributes to a lifestyle of not being aware of or prioritizing dental health in most African communities [31, 32, 33], and such, among other factors, could lead to cumulative burden of disease over the years which could be extremely profound in this population segment [3].

Self-rated dental health closely mimic actual clinical dental health status [34]. However, since dental health exists as a subset of oral health, which extends beyond dental outcomes to include the health of the entire oro-facial system [35], self-rated dental health is mostly studied alongside self-rated oral health with some surveys publishing it as self-rated oral health even if the variables studied only relate to teeth and gums [36].

The impact of dental disorders on self-rated dental health however remains understudied particularly in Kenya as in most other African countries [37]. This study assessed dental disorders and their impact on self-rated dental health of the elderly in select rural communities of Homa Bay County, Western Kenya.

## Materials and Methods

### Study Design and Setting

This was cross-sectional quantitative study conducted in North Karachuonyo ward of Homa Bay County. The County is situated in South-Western Kenya and is administered by eight sub-counties, 40 wards, 23 divisions, 140 locations and 265 sub-locations [38]. North Karachuonyo ward is situated within the GPS coordinates -**0.364015, 34.54282** and comprised four community administrative locations. The ward is characterized by poor access to improved water and sanitation with 99% of water sources being unimproved (mainly ponds) and 20% of the population practicing open defecation [39]. Educational levels are arguably suboptimal with 19% of the population having no formal education and majority (62.8%) having no more than primary education. Furthermore 18% of the population are either retired or homemakers and 7.4% have no gainful employment [39].

### Sampling

The sample size was determined using Yamane’s (1967) formula based on a study population of 1159, confidence level of 95%, and 5% precision. Participants were eligible for the study if they were aged 65 years and over, resided in the study area and gave consent to participate. Proportionate stratified sampling was used in order to ensure equitable sample distribution across all the four locations. The population was stratified by the four locations and the proportion to be included in the final sample (***i***) was then calculated using the formula:

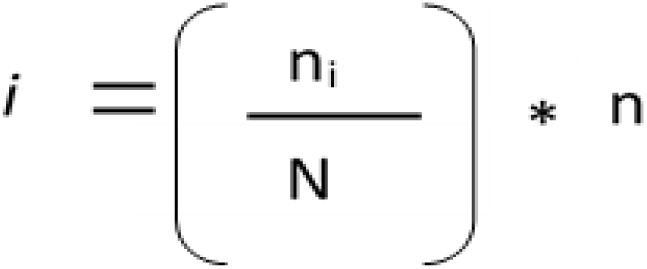

Where, n_i_ = number of eligible participants in the location,

N = total number of eligible participants in all the locations (i.e. study population)

n = the predetermined sample size (300)

This yielded a sub-sample distribution as indicated in table 1. Further to this simple random sampling was used to recruit participants.

**Table 1:** distribution of sample population across selected community administrative units.

| Community Unit | Proportion | Sample |
| --- | --- | --- |
| C1 | (163/1159) * 300 | 43 |
| C2 | (264/1159) * 300 | 68 |
| C3 | (237/1159) * 300 | 61 |
| C4 | (495/1159) * 300 | 128 |
| **Total** |  | **300** |

### Measurements

Data was collected between January and March 2020 through interviewer-administered questionnaires. The instrument collected data on socio-demographics, dental disorders and self-rated dental health. Sociodemographic data included age, gender, community unit, education, occupational history, household composition and self-rated economic status. Dental disorders assessed included self-reported dental pain, tooth extraction, tooth mobility, cavities gingival bleeding and dental trauma within the past one year. Self-rated dental health was assessed by a single question, “How would you rate your dental health?” with four response options; *very poor, poor*, *good*, *very good*. The instrument was initially drafted in English based on study objectives and a review of literature, translated into the local dialect (*Dholuo*), then back translated to English and pre-tested for reliability in a pilot study with 30 respondents. The instrument’s internal consistency reliability was assessed by computing Cronbach’s α coefficient for related constructs which yielded a value of 0.77. Those included in the pretest were excluded in the main study. Data was gathered through home visits with the help of trained research assistants who were community health workers.

### Ethics Statement

The study protocol was reviewed and approved by the Institutional Ethics Review Committee of the University of Eastern Africa, Baraton (IRB no. UEAB/REC/21/10/2019). In compliance with Kenyan standards, additional approval was granted by the Kenya National Commission for Science, Technology and Innovation (NACOSTI) as well as the Homa Bay County director of education. Written informed consent was obtained from all the study participants before enlisting them in the study. A written consent form was provided along with the questionnaire which the participants were obligated to read and accept or reject in witness of the research assistant. Those who were unable to read had the consent form read out to them by the research assistant. Only those who gave consent were included in the study. Furthermore, no physically invasive methods were employed in data collection as dental disorders were assessed through self-report.

### Data Analysis

Descriptive statistics including frequencies and percentages were used to summarize sociodemographic data as well as the distribution of dental disorders. Sociodemographic data was disaggregated by gender. Pearson’s chi-square test (*X^2^*) for independence was used to compare distributional differences with alpha set at p ≤ 0.05. Self-rated dental health was dichotomized into “good-very good/poor-very poor” from the original variable that had these four categories separately. The impact of dental disorders on self-rated dental health was estimated using the *phi* coefficient. Analyses were performed using IBM SPSS Statistics software version 26.0.

## Results

### Sociodemographic characteristics

Majority (64%) of the respondents were aged 65-74 years. Female respondents were more (55.3%) than males (44.75%). Most (56.7%) of the respondents had no more than primary education with about 3 in 10 (26.3%) having no formal education at all. There was statistically significant disparity in education with females constituting a majority (79%) of those without formal education and a minority (27%) of those with secondary education and above (p<0.00001). Further, 8 in 10 of the study sample never had any formal employment with nearly the same proportion (79.3%) self-rating their economic status as poor. Again there was statistically significant disparity in employment with females accounting for 60% of those that were never formally employed (p<0.0001). See table 1 for illustration.

### Prevalence of dental disorders

As indicated in the table below, Table 2, over three-quarters (76%) of the respondents had experienced dental pain within the last one year with dental trauma being the least occurring dental disorder in the study sample (16%). Nearly 7 out of 10 (67%) respondents had lost at least one tooth due to medical extraction whereas half of the respondents (52.3%) were having at least one carious tooth at the time of data collection. Periodontal symptoms were equally frequent in the study sample with 6 in 10 (60%) reporting gingival bleeding and nearly the same proportion (56%) reporting at least one mobile tooth.

**Table 2:** Sociodemographic characteristics of respondents.

| Characteristic | Frequency $n(\%)$ | Chi square ($p$-value) |
| --- | --- | --- |
| Gender |  | ref |
| Female | 166(55.3) |  |
| *Male* | 134(44.7) |  |
| Age |  | 3.775 (0.151) |
| 65-74 | 194(64.7) |  |
| 75-84 | 92(30.7) |  |
| 85 plus | 14(4.7) |  |
| Community unit |  | 5.124 (0.163) |
| C1 | 128(42.7) |  |
| C2 | 43(14.3) |  |
| C3 | 61(20.3) |  |
| C4 | 68(22.7) |  |
| Formal Education |  | 35.704 (<0.00001) * |
| None | 79(26.3) |  |
| Primary | 170(56.7) |  |
| Secondary plus | 51(17.0) |  |
| Occupational history |  | 16.467 (0.000049) |
| Never had formal employment | 247(82.3) |  |
| Was formally employed | 53(17.7) |  |
| Household composition |  | 7.751 (0.005) |
| Alone | 38(12.7) |  |
| With family | 262(87.3) |  |
| Economic Status |  | 7.125 (0.008) |
| Poor-very poor | 238(79.3) |  |
| Good-very good | 62(20.7) |  |

\*Exact *p*-value = 1.766E-8

### Relationship between dental disorders and self-rated dental health

As indicated in the table below, Table 3, over 6 in 10 (63.3%) of the respondents rated their dental health as poor. Chi-square test indicted there was statistically significant relationship between dental disorders and self-rated dental health among the study respondents. Judging by the test statistic associated *p*-values and corresponding *phi* coefficient, tooth extraction had the strongest impact on self-rated dental health (X^2^ =27.818, p<0.00001, φ = 0.305) followed by tooth mobility (X^2^ =27.180, *p*<0.00001, φ = 0.301), gingival bleeding (X^2^ =19.378, p=0.000011, φ = 0.254) and cavities (X^2^ =17.757, p=0.000025, φ = 0.243). Respondents’ experience of dental trauma (X^2^ = 9.843, p= 0.002, φ = 0.181) and dental pain (X^2^ = 8.842, p= 0.003, φ = 0.172), and the degree of dental discoloration (X^2^ =10.992, p=0.001, φ = 0.194) had the least impact on self-rated dental health.

**Table 3:** prevalence of dental disorders (frequency and percentages)

| Disorder | Frequency<br>(n) | Percent<br>(%) |
| --- | --- | --- |
| Dental Pain |  |  |
| *No* | 72 | 24.0 |
| *Yes* | 228 | 76.0 |
| Tooth extraction (medical) |  |  |
| *No* | 99 | 33.0 |
| *Yes* | 201 | 67.0 |
| Cavities |  |  |
| *No* | 143 | 47.7 |
| *Yes* | 157 | 52.3 |
| Gingival Bleeding |  |  |
| *No* | 120 | 40.0 |
| *Yes* | 180 | 60.0 |
| Tooth Mobility |  |  |
| *No* | 132 | 44.0 |
| *Yes* | 168 | 56.0 |
| Dental trauma |  |  |
| *No* | 252 | 84.0 |
| *Yes* | 48 | 16.0 |

**Table 4:** impact of dental disorders on self-rated dental health.

| | Self-rated dental health | | $X^2$ (p-value) | Phi coeff |
| --- | --- | --- | --- | --- |
| *N* = 300 | *good*<br>(36.7%) | *poor* (63.3%) |  |  |
| Dental Pain |  |  | 8.842 (0.003) | 0.172 |
| *no* | 37(51.4)^ | 35(48.6) |  |  |
| *yes* | 73(32.0) | 155(68.0) |  |  |
| Cavities |  |  | 17.757 (0.000025) | 0.243 |
| *no* | 70 (49.0) | 73 (51.0) |  |  |
| *yes* | 40 (25.5) | 117 (74.5) |  |  |
| Gingival bleeding |  |  | 19.378 (0.000011) | 0.254 |
| *no* | 62(51.7) | 58(48.3) |  |  |
| *yes* | 48(26.7) | 132(73.3) |  |  |
| Tooth mobility |  |  | 27.180<br>(<0.00001)* | 0.301 |
| *no* | 70(53.0) | 62(47.0) |  |  |
| *yes* | 40(23.8) | 128(76.2) |  |  |
| Dental trauma |  |  | 9.843 (0.002) | 0.181 |
| *no* | 102(40.5) | 150(59.5) |  |  |
| *yes* | 8(16.7) | 40(83.3) |  |  |
| Tooth extraction (medical) |  |  | 27.818<br>(<0.00001)** | 0.305 |
| *no* | 57(57.6) | 42(42.4) |  |  |
| *yes* | 53(26.4) | 148(73.6) |  |  |
| Discoloration |  |  | 10.992(0.001) | 0.194 |
| *slight* | 68(46.9) | 77(53.1) |  |  |
| *very* | 41(28.1) | 105(71.9) |  |  |

\*exact *p*-value = 1.8541E-7
\*\*exact *p*-value =1.3328E-7
^the row percentages are within categories of dental disorders

## Discussion

Dental health is critical to general health and wellbeing of geriatric population, as it is considered a basic human right and recognized as essential tenet in public health and a core indicator of general health and quality of life [40, 41]. Notwithstanding, dental disorders remain a leading geriatric disease in Kenya and other developing countries, compromising their self-expression, nutrition and general psychological and social wellbeing [24, 42, 43].

This study investigated the impact of dental health disorders on self-rated dental health among the elderly in parts of rural Kenya. The findings of this study showed that there was a high prevalence of dental disorders among the elderly with dental pain having been experienced by nearly 8 in 10 (76%). Compared to other studies, the prevalence of dental pain in this study was higher than those reported among the elderly in Hong Kong (62%) [44], India (39%) [45], Brazil (32%) [24] and a low of 19% in South Africa [46].

Notably, reported tooth mobility prevalence in this study (56.0%) was comparable to those reported in the US (56.5%) but lower than those reported in parts of Africa (91.0%), while the prevalence of gingival bleeding (60.0%) was higher than those reported among the elderly in parts of China (57.8%) and Egypt (52%) [11-12, 47-48]. In literature, these conditions are a hallmark of periodontal disease which is among the leading dental disorder among the elderly [47, 48].

Furthermore, the prevalence of cavities in this study (52.3%) was higher than those reported in parts of India (41.9%); Malawi (49.0%) and the US (47.0%) [9, 10, 13], but lower than reported in parts of china (67.5%) [47]. Again, this study found that nearly 7 in 10 (67%) had lost at least a tooth to medical extraction. In literature, medical tooth extraction is generally attributed to untreated dental caries and periodontal disease [50-52]. This finding therefore indicate that the elderly in the study population could have endured or neglected these conditions and only visited dentists too late in the natural history of the diseases. Some researchers have argued that such prevalence of dental disorders indicate that large proportion of the elderly either lack access to or neglect to utilize effective oral healthcare programs [53].

Self-rated dental health closely mimic actual dental health status and is recommended for socio-dental assessment particularly among the elderly in low-resource settings [54]. However, since dental health exists as a subset of oral health, which extends beyond dental outcomes to include the health of the entire oro-facial system [35], self-rated dental health is mostly studied alongside self-rated oral health with some surveys publishing it as self-rated oral health even if the variables studied only relate to teeth and gums [36]. Previous studies have validated the use of this technique in various settings and study populations [54]. Lundbeck and others, for instance, conducted a study in 2020 among elderly New Zealanders to determine the clinical validity of self-rated oral health. The study concluded that self-rated oral health accurately depicted the clinical oral health status [55].

In this study, more than 6 in 10 (63.3%) respondents self-rated their dental health as poor. This was in stark contrast to findings in some higher income countries such as United States (28%) [53], Japan (40%) [56] and Kingdom of Saudi Arabia (24%) [57] but comparable to Thailand (67%) [54]. This is likely because of better access to oral care resulting in better oral health outcomes among the elderly in HICs compared to middle income countries. Studies have consistently reported better general and oral health outcomes in HICs.

Notwithstanding, Locker and two others argued in a landmark study on self-ratings of oral health that other frames of reference impact self-rated oral health in addition to current oral problems [58]. The authors posited that such factors as respondent’s emotional or physical state, their demographic characteristics, socioeconomic status, underlying diseases, as well as health-related behaviors are all frames of reference and could impact their self-rating of oral health. This study therefore undertook to establish the specific impact that dental disorders have on self-rated dental health. The findings revealed that dental disorders disproportionately but significantly impacted respondents’ self-rating of oral health status. The overall impact of dental disorders on respondents’ self-rated oral health however appeared quite marginal considering the corresponding effect size, but we cannot make any conclusive assertions on this considering the scope and design of this study.

Medically driven tooth loss had the strongest impact on self-rated dental health (φ = 0.305, *p*<0.00001) followed by tooth mobility (φ = 0.301, *p*<0.00001), gingival bleeding (φ = 0.254, *p*=0.000011) and cavities (φ = 0.243, *p*=0.000025), whereas respondents’ experience of dental trauma (φ = 0.181, *p*=0.002) and dental pain (φ = 0.172, *p*=0.003) had the least impact on self-rated dental health. According to the interpretational principles for φ espoused by Akoglu (2018) [59], tooth extraction and tooth mobility had strong impact on self-rated dental health, with the rest having moderate to high impact. These findings suggest that the elderly in rural communities in Kenya value tooth retention therefore any loss of teeth, particularly on medical grounds, strongly impact their perception of oral wellbeing. This interpretation is further supported by the finding that tooth mobility came a close second to tooth loss on impact in the sense that the participants could have perceived any mobile tooth as a step toward tooth loss.

Furthermore, respondents’ degree of dental discoloration also had an impact on their self-rated dental health (φ = 0.194, p=0.001) suggesting that even dental aesthetics was a concern among the studied population. This finding contrasts the findings of a study conducted in Nigeria [60] which concluded that dental discoloration and the desire to medically treat it was only significant among the younger age-groups. Our findings suggest that the African elderly population is mindful of their tooth color and probably just lack access to dental aesthetics.

This study was not without limitations. First, since the design was cross-sectional, it was not possible to establish cause-effect relationship. Secondly, dental disorders were assessed through self-report which, though an established methodology, is not as robust as clinical examination. Lastly, this study focused on the bivariate relationship between explanatory and outcome measures therefore the impact of any possible extraneous variables was not explored. Nonetheless this study’s findings offer insightful evidence as regards the differential impact of dental disorders on self-rated dental health among older adults in rural Kenya and is to the best of our knowledge the first of its kind.

## Conclusion

This study investigated the impact of dental disorders on self-rated dental health among the elderly in rural communities in Kenya. The study established that dental disorders significantly but differentially impacted self-rated dental health of the elderly, with tooth loss and tooth mobility being the leading disorders in impact. Addressing the causes of these disorders among the elderly in Kenya is critical to improving oral health and wellbeing.

## Author Contributions

W.A—conceptualized the study and significantly worked on the manuscript, revised, and provided approval for submission. F.N.-manuscript preparation, and revisions on all drafts. All authors have read and agreed to the published version of the manuscript.

## Institutional Review Board Statement

The study was conducted according to the guidelines of the Declaration of Helsinki and approved by the Ethics Committee of the University of Eastern Africa, Baraton (IRB no. UEAB/REC/21/10/2019).

## Informed Consent Statement

Informed consent was obtained from all subjects involved in the study by signing a consent form that had explanation on the voluntary participation and purpose of the study explained to them.

## Data Availability Statement

The datasets used and/or analyzed during the current study are contained within this article. Any additional data or clarification available upon reasonable request.

## Acknowledgments

We are grateful to all the research assistants and field coordinators who helped with the study. We acknowledge participation of those who accepted to be interviewed for their time and shared experiences with us.

